# Reducing delays in seeking abortion among women who visit health facilities for safe abortion service in Mekelle City, Ethiopia: Pre-post interventional study

**DOI:** 10.1101/2022.10.18.22281207

**Authors:** Awol Yemane Legesse, Mengistu Welday, Mengistu Hagazi, Mussie Alemayehu, Mache Tsadik, Ambachew Hailemicheal, Beyene Meressa, Daniel Gebre, Hale Teka

**Affiliations:** Department of Obstetrics and Gynecology, College of Health Sciences, Mekelle University, Mekelle, Tigray, Ethiopia; Department of Midwifery, College of Health Sciences, Mekelle University, Mekelle, Tigray, Ethiopia; Department of Health Systems, College of Health Sciences, Mekelle UniversityMekelle, Tigray, Ethiopia; Department of Reproductive Health, College of Health Sciences, Mekelle University. Mekelle, Tigray, Ethiopia; Tigray Regional Health Bureau, Mekelle, Tigray, Ethiopia; Department of Public Health, College of Health Sciences, Mekelle University, Mekelle, Tigray, Ethiopia; Labor ward, Ayder Comprehensive Specialized Hospital, Mekelle, Tigray, Ethiopia

**Keywords:** Abortion, Pregnancy termination, Ayder, Health education, Tigray region, Ethiopia

## Abstract

**Objectives:** **T**his study assessed the effect of health education on reducing the delay in seeking abortion in Mekelle health facilities, Tigray region, Ethiopia.

**Design:** A pre-post interventional study design with sample size of 322 women.

**Setting:** The study was conducted in 12 health facilities at Mekelle, Tigray, Ethiopia between February 1^st^ through September 30^th^, 2020

**Participants:** All women who have attended safe abortion services in Mekelle health facilities during the time of the data collection period were the study population.

**Intervention:** Women education on the Ethiopian abortion law, availability of abortion services, and optimal time of pregnancy termination using 1000 leaflets, 20 street posters for 3 months.

**Results:** Compared to the pre-intervention group, a much change in reducing the gestational age in weeks was observed in the post intervention period to terminate the pregnancy with 9.8 decrease per 100 respondents (95% CI 9.25 to 10.36) and a Cohen’s d value of 5.23 was found. Besides, there was statistically significant difference between the pre and post-intervention on the respondent knowledge on the possibility of termination pregnancy based on wish according to Ethiopian abortion law at a p-value of < 0.024.

**Conclusions:** Women’s health education on the availability of safe service and optimal gestational age for termination has led to a significant reduction of the delay in weeks with large effect size. Moreover, it brings a statistically significant difference on respondent knowledge of Ethiopian abortion law. We recommend further randomized contrail study involving three face delays of safe abortion service.

## Introduction

Abortion is the termination of pregnancy before fetal viability, which is conventionally taken to be less than 20 weeks from the Last Normal Menstrual Period (LNMP). If the LNMP is not known a birth weight of less than 500 gm is considered an abortion.^1^ According to Ethiopia Abortion law, it is considered to be less than 28 weeks or less than 1000 gm.^2^ Safe abortion is when it is performed by people with the necessary skills or using an appropriate technique, and/or in an environment that meets minimum medical standards.^3^ First Trimester abortion is termination of pregnancy before 13 weeks. Second trimester abortion is the termination of pregnancy in a period from 13 to 28 weeks of gestation, which again is subdivided into early period between 13 and 20 weeks and late period between 20 and 28 weeks.^4, 5^

Despite of the majority of abortions are performed in the first trimester, 10–15% of abortions have been taking place in the second trimester. In developing countries, compared with first-trimester abortion, second-trimester abortion has increased contribution to maternal morbidity and mortality. The ill effect of second-trimester abortion is especially immense in developing countries where access to safe second-trimester abortion is limited.^6^ In sub-Saharan Africa(SSA), 2/3^rd^ of the total unsafe abortion complications, were reported to be during the second trimester period. The risk of death is 75 times higher among those women who had an abortion in their second trimester of pregnancy than those women who had in their first trimester.^7^ In Ethiopia, about half of all health facilities provide safe induced abortion services. Whereas access to second-trimester abortion services is severely limited. ^8^

To this end, women in the second trimester tend to visit unsafe abortion services to get their pregnancy terminated. According to a study done in Jimma and Arsi, most abortions occur in the young vulnerable age group.^9,10^ Studies done after the revision of abortion law have shown the occurrence of unsafe abortions in the ground. Most of the unsafe abortions occur in women with less educational status and less access to health care facilities.^12^ Therefore, interventional studies including ours can be applied at the community level to reduce the delay of gestational age in weeks to terminate the pregnancy by providing health education to the target group.

Thus, the rationale of this study was assessing the provision of health education using street poster and client information sheet would be possible to reduce the delay in seeking abortion service and increase women’s knowledge on the possibility of termination of pregnancy based on a certain circumstance. Hence, we carried out a pre post study that aimed at addressing the delay in seeking safe abortion services in Ethiopia in general and in Tigray region in particular.

## Material and Methods

### Study area and period

The study was conducted in Mekelle City Health facilities, Tigray Region, Ethiopia between February 1^st^ through September 30^th^, 2020 among women who presented for safe abortion services. Mekelle city is the capital city of the Tigray region and is found 783 km in the north away from the capital city of Ethiopia, Addis Ababa. It has a population of 233,012, out of this 119,765(51.3%) are Females. It has 7 kifle-ketema (sub city), 72 ketena and 54,073 households. In Mekelle, there are 2 public hospitals, 10 health centers making the health service coverage 56%.

### Study Design and source population

A pre-post interventional study design was conducted to answer the objective of the study. All women who have attended safe abortion services in Mekelle health facilities during the time of the data collection period were the study population whereas those who declared ill during data collection and women who gestational trophoblastic disease (partial mole) were excluded from the study.

### Sample size determination

The sample size was determined based on the double population proportion formula using open epi info software. The assumptions made were: two-sided significance level 95, power 80%, unexposed /Exposed 1, unexposed with outcome 15 %, ^13^ exposed with outcome 5%. The total minimum sample size calculated was 322(161 for pre and 161 for post-intervention). Stratified consecutive sampling was employed. Health facilities were stratified and individuals were selected by consecutive sampling techniques. The health facilities for second-trimester abortion were Ayder Comprehensive Specialized Hospital (ACSH) and Mekelle general hospital. For the first trimester, all health centers and private clinics in Mekelle City were included. The stratification was made based on case flow of the health facilities.

### Description of the intervention

The intervention was developed based on reviewing different literatures and discussion with service providers. From literature, the study employed a three-face delay model from pieces of literature focusing on the first delay. That is a delay in decision-making to get a pregnancy-termination service. Along with such information was strengthen by making discussion with health care provider. The intervention aimed to decrease the time gap between pregnancy diagnosis and the decision to terminate the pregnancy. Hence, its goal was to decrease the proportion of women transitioning to second trimester. Accordingly, the following intervention (using street poster and client information sheet) was done on the ground. The algorism followed for the intervention was described below.

1. Is the pregnancy planned, wanted, and supported? If yes visit a health facility for an early ultrasound and antenatal care If not, you should visit a health facility for an early ultrasound and counseling
2. Is the pregnancy the result of incest, rape, or underage?
3. If yes, do you have an intention to terminate the pregnancy? Safe abortion service is provided at all governmental health centers for free. If you want to terminate the pregnancy, the earlier the gestational age the safer. Later gestational age termination of pregnancy is associated with morbidity and mortality. The intervention was monitored through supervision, whether the vendors are providing the leaflets by making random checks at 10% of the facilities (Figure 1).

**Figure 1:**
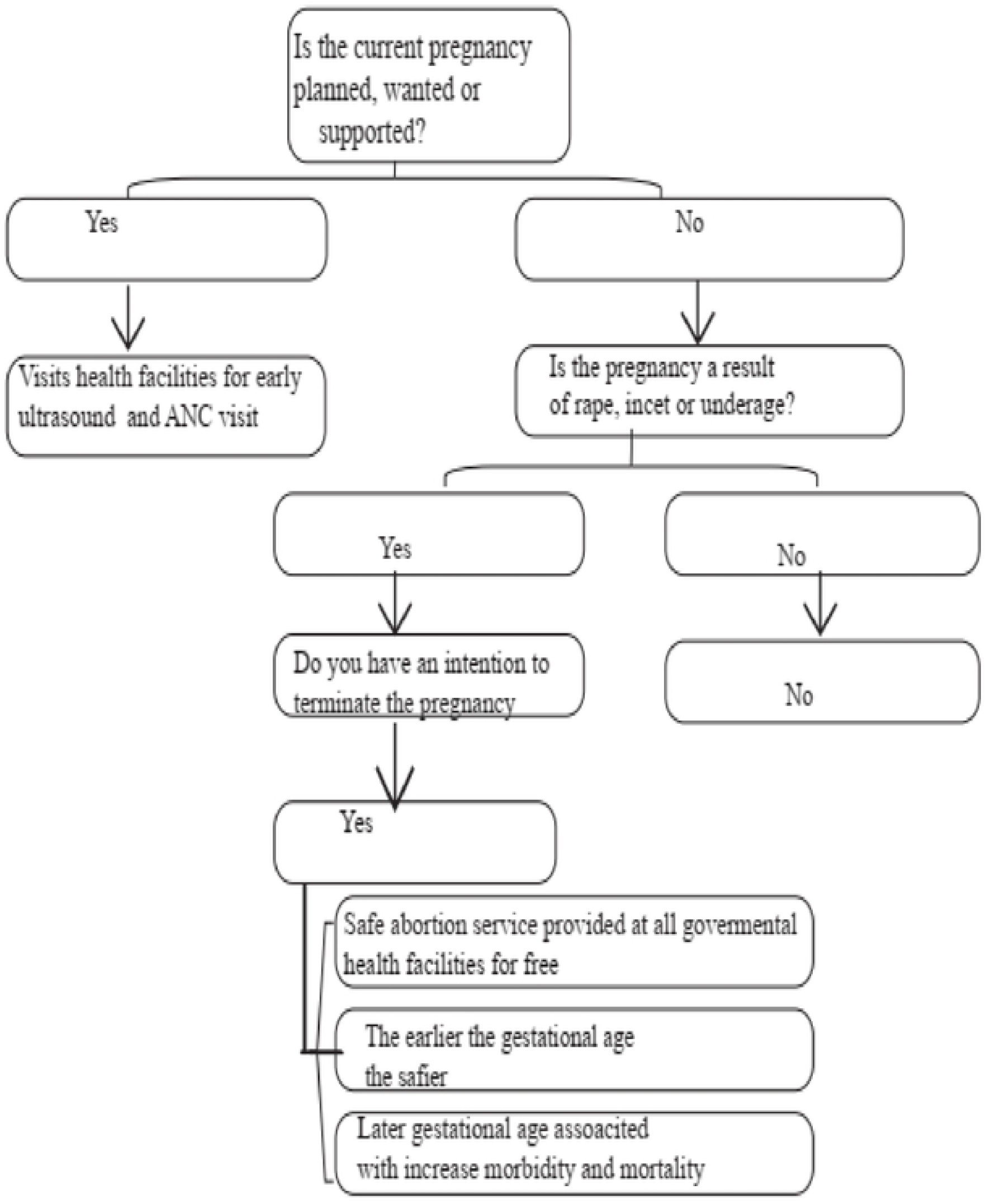
Intervention description of the pre and post-intervention abortion study in health facilities of Mekelle City, Tigray region, Ethiopia, 2020

The above listed algorism was done using street poster and client information sheet. The detail description of each intervention is described below.

- A street posters were distributed and used at diagnostic clinics, drug vendors, and health centers. A street poster was distributed to diagnostic clinics, drug vendors, and health centers. These areas are usually where women come for a pregnancy checkup. For this purpose, a total of 20 posters were distributed.
- A client information sheet was provided to the client. For this purpose, a total of 1000 client information sheet was given to 10 health centers, 30 pharmacies, 15 laboratories. The information leaflet was provided to the clients when they present for a pregnancy test at pharmacies, laboratories, and health centers.

### Data collection

A structured questionnaire was used to collect the data. The questions and statements were assessed, grouped, and arranged into socio-demographic and economic conditions(age, marital status, residence, occupation, educational status, ethnicity and average monthly income), reproductive health issues(nature of menses, gravidity, parity, number of live births, current conditions of pregnancy, contraceptive history, and abortion history), contraceptive uses, and other factors related to maternal health like taking time while finding money, transportation problem, referral problem, and distant from the institute were collected. After extensive revision, the final version of the English questionnaire was developed. An individual who had a very good ability of both English and Tigrigna languages translated the English version and another individual of similar ability then translated the agreed Tigrigna version of the questionnaire back to English to check for any inconsistencies or distortion in the meaning of words. An information technology person developed the open data kit (ODK) template. The paper-developed questionnaire was exported to ODK. Hence, the pre intervention (baseline) and post-intervention (end-line) data were collected using ODK with three months duration. The data collected via ODK was sent to the Mekelle University server. Ten midwives were used for the data collection. Intensive training was given to the data collectors and supervisors. Regular supervision and continuous follow-up were made by the supervisors The paper-based items were transferred to the mobile-based application (ODK) which ensures skip pattern; immediate scanning of the collected tool in the server, friendly to use, and avoids cost for paper duplication. The progress of the intervention was monitored by research team members in addition to the supervisor.

### Outcome variables

The primary outcome for the study was reducing delay in seeking abortion service whereas respondent’s knowledge on termination of pregnancy in accordance with the national guideline

### Measurement

#### Second trimester abortion

abortion after gestational age of 11 weeks plus six days. It was measured by asking the women gestational age and confirmed the result using ultrasound.

#### Gestational age

The number of days or weeks since the first day of the woman’s last normal menstrual period (LMP) dating by ultrasound.

Delay in seeking abortion: if a woman seeks an abortion after one week of pregnancy diagnosis and or if she presents during second-trimester pregnancy.

#### Women’s and partners’ educational status

It was categorized in to **‘**No education’, ‘Primary education’, ‘Secondary education’ and ‘Higher’ respectively for respondents who were not able to read and write a simple sentence, 1-8^th^ grade, 9-12^th^grade and college and university level.

#### Marital status

It was categorized as “*single*” for women who were single and unmarried, “*married*” for those married women and living together and “*other*” for widowed and divorced. **Knowledge of respondents on circumstances to terminate a pregnancy**. It was collected with question of “*According to Ethiopian law, is it possible to terminate pregnancy based on wish”*. And, the response was categorized as 1 “*Yes*” and 2 “*No*”. However, in Ethiopia abortion was legalized on the circumstance of the following points. These were if the pregnancy was as result of *rape, incest, lethal congenital* anomalies *of the fetus, mother has physical or mental disability and “under age pregnancy*”. Hence, women who respond “No” to the above question were labeled as “have knowledge on circumstance to terminate a pregnancy” while those who respond “*Yes*” were labeled as “don’t have knowledge on circumstance to terminate a pregnancy”.

### Data quality control

Data collectors and supervisors were trained adequately and regular supervision was made in place throughout the data collection period of both pre intervention and post intervention period. To ensure a complete and consistence data collection, a regular check-up was made in place in the field on daily basis. The data were collected using a mobile based application that ensures a skip pattern, it reduce cost for data collection reduces error and time during paper printing and data entry, user friendly, allows control of the allotted time for a questionnaire, and enables the immediate transfer of collected data to the server.

### Data Analyses

The data collected using ODK, was exported to SPSS version 23 for analysis. At the beginning, descriptive statistical analysis was carried out to describe the distribution of the data across the selected factors related to dependent variable (reducing gestational age in weeks and knowledge of the respondents on the termination of pregnancy according to the national guideline). For categorical variables, frequencies and percentages were calculated while mean (SD) was calculated for normally distributed continuous variables.

To analysis the change in gestational age in weeks using ultrasound to terminate the pregnancy an independent t-test was carried out. The result was described using t-test, df, P-value, mean value of both groups (pre intervention and post intervention and adjusted risk with its 95% CI. Along with, Cohens’d was collected to determine the effect size of the intervention. It was calculated based on comparing two means with value ranging from 0(“no difference”) to infinity. A value of d=2,” small”, d=0.5=“ medium” and d=0.8” large” indicates the strength of the effect size. A *“d”* of three indicates the two groups differ by three standard deviations, and so on.^14^

Along with, McNemar’s test was carried out to determine if there were differences on a women’s knowledge on the possibility of termination pregnancy based on Ethiopia law for women who attended safe abortion services to health facilities. Before running the test, assumption of the test was done. The assumptions were the dependent variable must be a categorical variable with two categories (i.e. a dichotomous variable), the groups of the dependent variable must be mutually exclusive (no groups can overlap) and the respondents are a random sample from the population interest. Hence, the McNemar’s test was carried once the assumption was fulfilled.

### Ethics approval and consent to participate

The study protocol was approved by the Institutional Review Board (IRB) of Mekelle University College of Health Sciences with a reference number of ERC-13159/2019. Permission to carry out the study in the Mekelle city was sought from Tigray Regional Health Bureau (TRHB) and Mekelle City Health Office. Informed written consent from each of the respondents before conducting the interviews was obtained. An information sheet that includes the purpose of the study, possible benefits and risk from enrolling the study, the right of the individual to withdraw from the study at any time of data collection, and the voluntary nature of participation was attached to each questionnaire for this purpose. In addition, the privacy and confidentiality of the collected information and arrangement for the identification of informants only through specific identification numbers was included in the information sheet. The authors authors did not have access to information that could identify individual participants during or after data collection.

### Patient and public involvement

Patients and/or the public were not involved in the design, or conduct, or reporting, or dissemination plans of this research.

## Results

### Socio-demographic characteristics of respondents

In both (pre and post intervention), a total of 166 pregnant women seeking abortion services were enrolled for each data collection time. The gestational age of all the cases was confirmed with ultrasound. The overall mean age of respondents in the pre intervention was 23.2 (±4.7) years, while it was 25.1(±4.5) years in the post intervention. The mean age of respondents for the pre and post-intervention group was 23.2 (±4.7) and 25.1(±4.5) years, respectively. Seventy two percent of the respondents in the pre intervention and 60% in the post intervention were single in marital status. In both (pre and post intervention) data, in average six-in-ten of the respondents were secondary in educational status. Moreover, one-in-four (25.9%) of the respondents in the pre-intervention and 36.7% in the post-intervention were employed in occupation status (Table 1).

**Table 1.** Socio-demographic profile of respondents in pre and post-intervention abortion study in health facilities of Mekelle City, Tigray region, Ethiopia, 2020.

| **Variable** | **Pre-intervention** | **Post intervention** | **P-value** |
| --- | --- | --- | --- |
|  | n(%) | n (%) |  |
| **Age** |  |  |  |
| 15-24 | 111(66.9) | 78(47) | 0.00001 |
| 25 and above | 55(33.1) | 88(53) | 0.0002 |
| Mean age (SD) | 23.2 ($\pm 4.7$) | 25.1($\pm 4.5$) | |
| **Marital status** |  |  |  |
| Single | 120(72.3) | 99(59.6) | 0.015 |
| Married | 28(16.9) | 40(24.1) | 0.102 |
| Others*(divorced, widowed) | 18(10.8) | 27(16.3) | 0.149 |
| **Educational status** |  |  |  |
| No formal education | 5(3) | 7(4.2) | 0.55 |
| Primary | 14(8.4) | 21(12.7) | 0.21 |
| Secondary | 100(60.2) | 93(56) | 0.43 |
| More than secondary | 47(28.3) | 45(27.1) | 0.80 |
| **Occupational status** |  |  |  |
| Student | 40(24.1) | 42(25.3) | 0.79 |
| Unemployed | 21(12.7) | 34(20.5) | 0.054 |
| Employed | 43(25.9) | 61(36.7) | 0.033 |
| Daily laborer | 33(19.9) | 21(12.7) | 0.07 |
| Housewife | 26(15.7) | 6(3.6) | 0.0001 |
| Others** (CSW) | 3(1.8) | 2(1.2) | 0.6523 |
| **Partner educational status** |  |  |  |
| No formal education | 2(1.2) | 3(2.3) | 0.65 |
| Primary | 3(1.8) | 1(0.8) | 0.65 |
| Secondary | 43(25.9) | 64(48.5) | 0.013 |
| More than secondary | 21(12.7) | 64(48.5) | 0.00001 |
| **Address** |  |  |  |
| Mekelle | 142(85.5) | 157(94.6) | 0.005 |
| Out of Mekelle | 24(14.5) | 9(5.4) | 0.0059 |

\*Widowed and divorced, \*\*Commercial sex workers

### Reproductive history of respondents in the pre and post-intervention period

On average, 65.1% of the respondents in the pre-intervention and 50.6% in the post intervention were prim gravida. The proportion of respondents who were nulliparous was 64.5% in the pre-intervention while it was 48.6% in the post-intervention. Moreover, 33.7% of the respondents in the pre-intervention had one and above alive children while it was increased to 42.2% in post-intervention. The figure of those respondents who had abortion was 3% in the pre-intervention whereas it was 11.4% in post intervention. Majority of the respondents in both data had unplanned pregnancy which accounts for 97.6%. The use of contraceptive prior the last pregnancy was increased from 51.8% in the pre-intervention to 85.2% in the post-intervention. Based on the post-intervention data, the most common type of contraceptive used prior the last pregnancy was emergency contraceptive that accounts for 60.9% (Table 2).

**Table 2.**
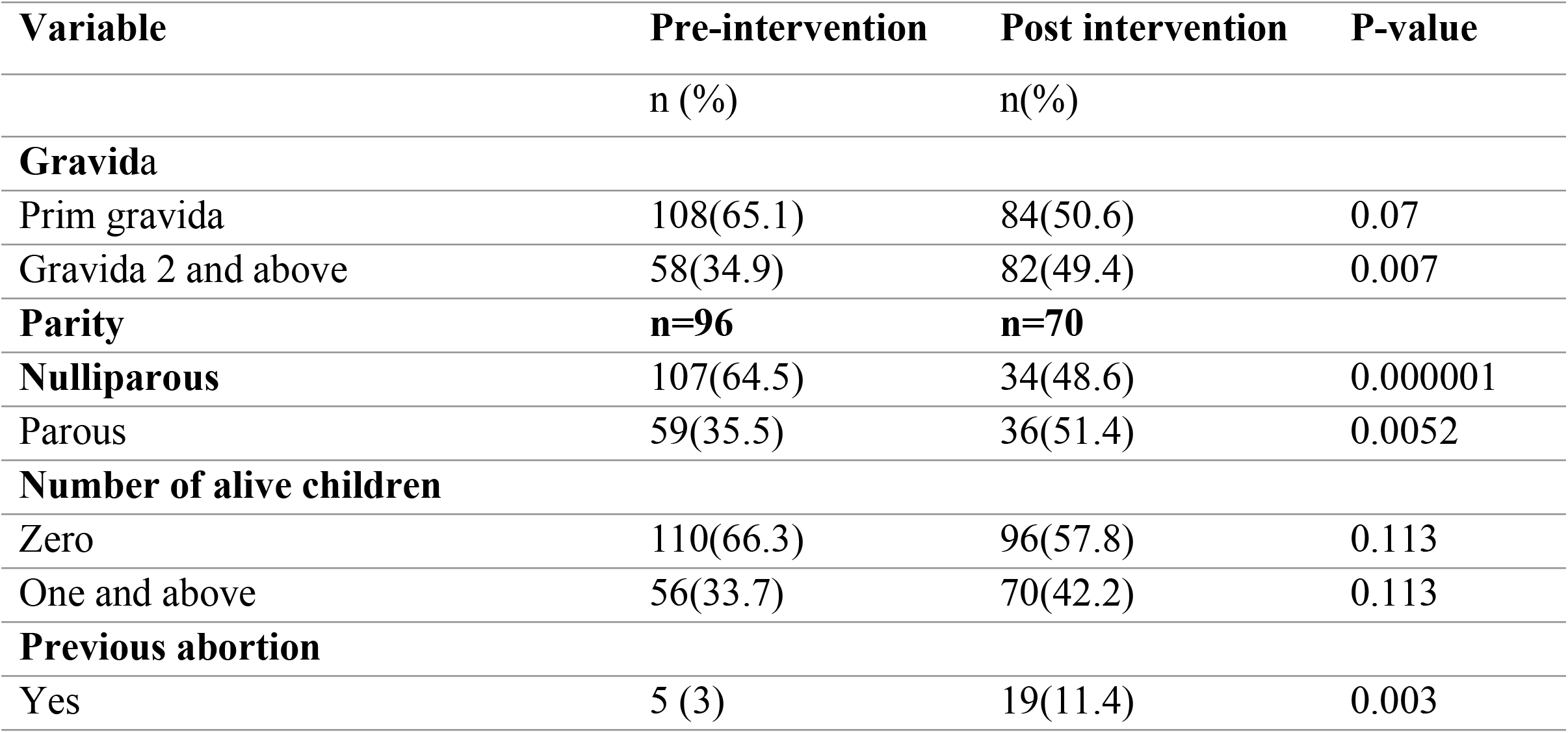
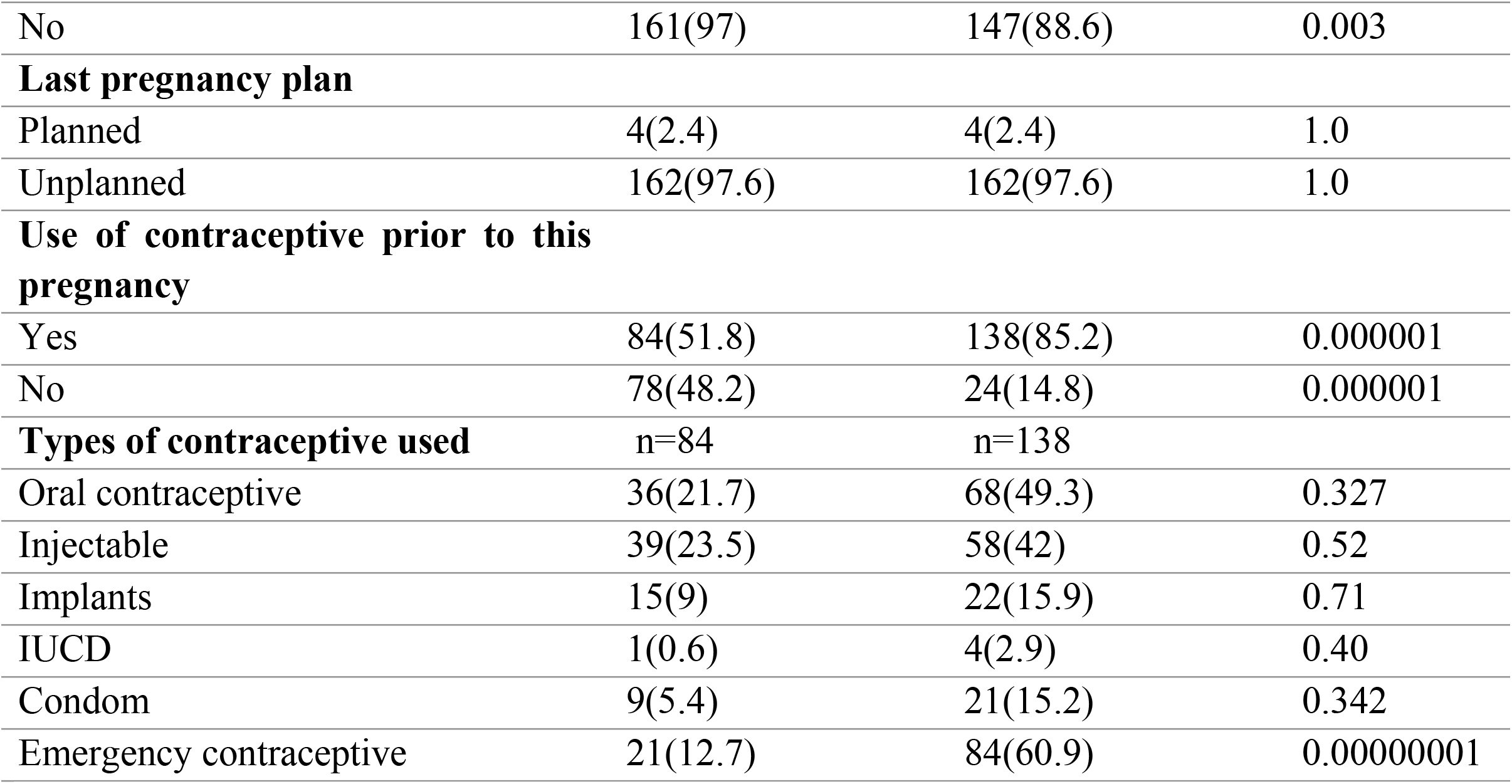
Reproductive history of respondents in pre and post-intervention in health facilities of Mekelle City, Tigray region, Ethiopia, 2020.

### Abortion related factors of respondents in the pre and post intervention period

The figure for the proportion of women who knew about their LNMP was increased from 44% in the pre-intervention to 80.7% in the post intervention. Along with, the mean gestational age was 12.12(±6.23) and 7.5(±2.33) weeks for pre-intervention and post-intervention respectively. Vomiting was the commonest symptom of the last pregnancy accounted for 87.9% in the pre-intervention and 95.6% in the post intervention. In both intervention data, 83.1% of the respondents reported their last pregnancy was confirmed in health facilities. The proportion of women who wish to terminate their last pregnancy earlier was decreased from 85.5% in the pre-intervention to 65.15% in the post-intervention. Along with, 69.3% of the respondents in the pre-intervention and 97.6% in the post-intervention data they decided immediately to make decision after confirmation of their last pregnancy. In average, the period between the decision to terminate the pregnancy and actual termination of the current pregnancy was 3.57(±8.6)weeks in the pre-intervention and 1.82(±3.76) weeks in the post intervention. Lack of money was mentioned by 24.7% in the pre-intervention and 65.7% in the post-intervention as a main reason for delay to seek abortion care after decision to terminate the pregnancy (Table 3).

**Table 3:**
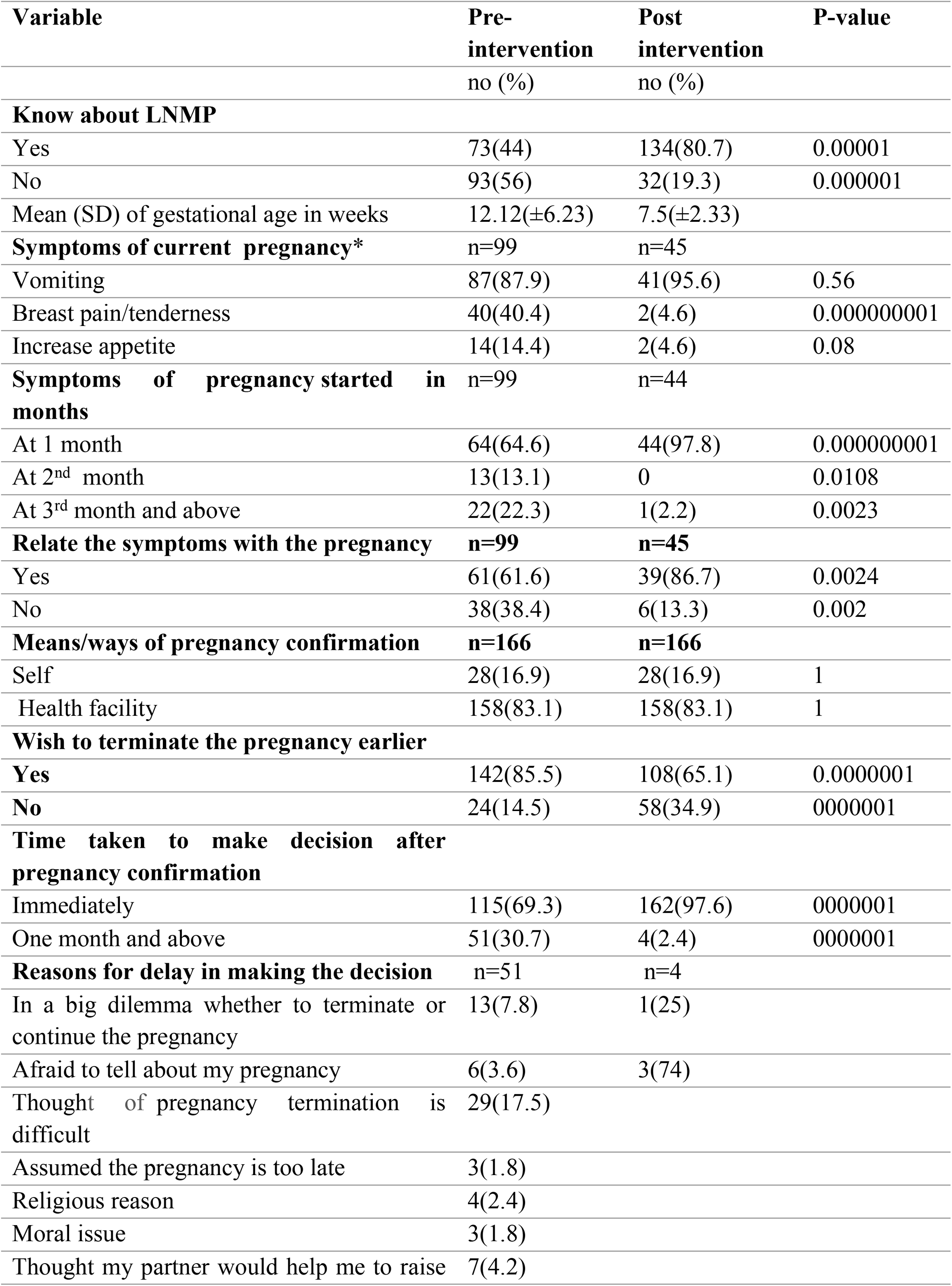
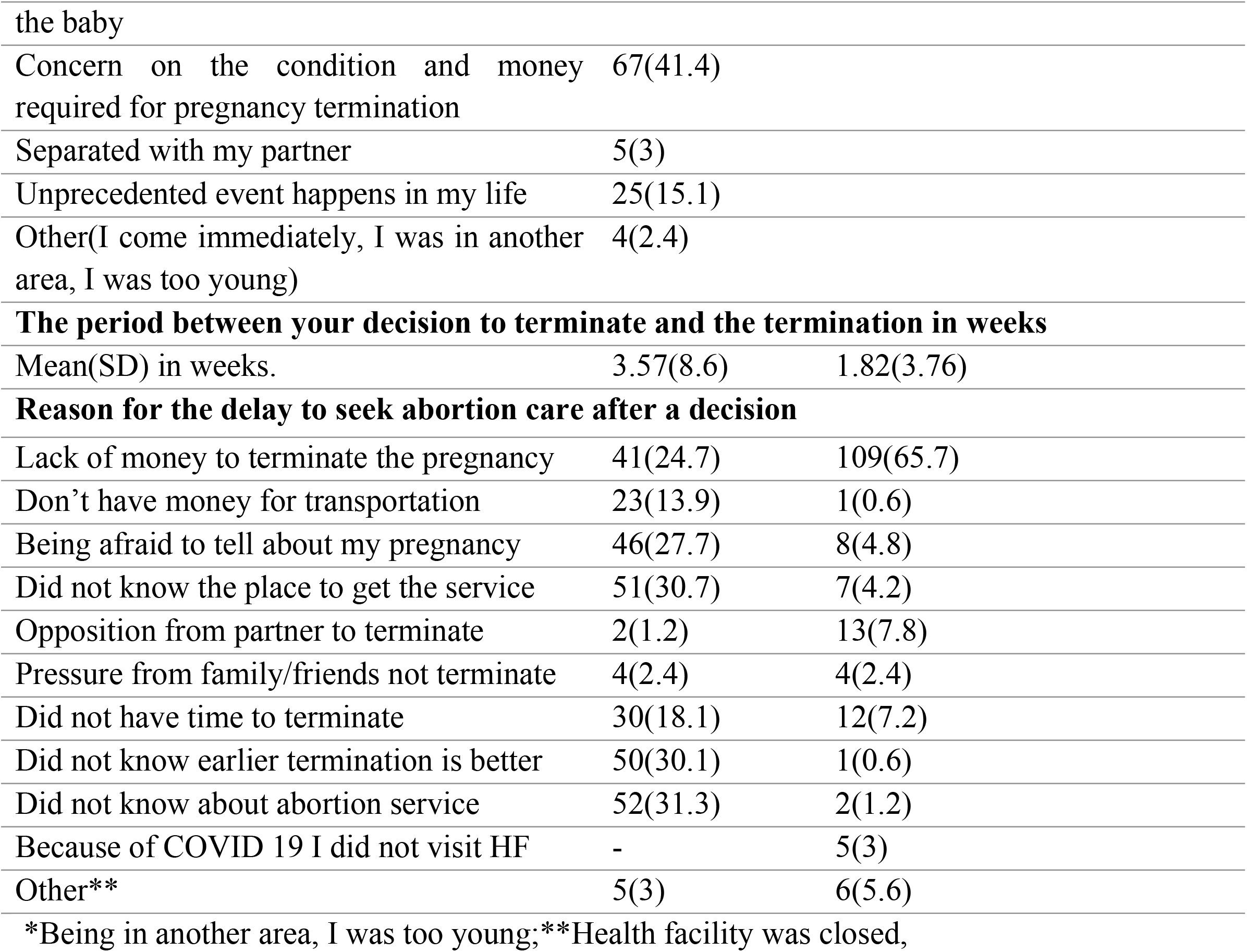
Abortion and pregnancy characteristics of clients seeking abortion in health facilities of Mekelle City, Tigray region, Ethiopia, 2020.

| Variable | Pre-intervention<br>no (%) | Post<br>intervention<br>no (%) | P-value |
| --- | --- | --- | --- |
| **Know about LNMP** |  |  |  |
| Yes | 73(44) | 134(80.7) | 0.00001 |
| No | 93(56) | 32(19.3) | 0.000001 |
| Mean (SD) of gestational age in weeks | 12.12(±6.23) | 7.5(±2.33) | |
| **Symptoms of current pregnancy\*** | n=99 | n=45 |  |
| Vomiting | 87(87.9) | 41(95.6) | 0.56 |
| Breast pain/tenderness | 40(40.4) | 2(4.6) | 0.000000001 |
| Increase appetite | 14(14.4) | 2(4.6) | 0.08 |
| **Symptoms of pregnancy started in months** | n=99 | n=44 |  |
| At 1 month | 64(64.6) | 44(97.8) | 0.000000001 |
| At 2<sup>nd</sup> month | 13(13.1) | 0 | 0.0108 |
| At 3<sup>rd</sup> month and above | 22(22.3) | 1(2.2) | 0.0023 |
| **Relate the symptoms with the pregnancy** | **n=99** | **n=45** |  |
| Yes | 61(61.6) | 39(86.7) | 0.0024 |
| No | 38(38.4) | 6(13.3) | 0.002 |
| **Means/ways of pregnancy confirmation** | **n=166** | **n=166** |  |
| Self | 28(16.9) | 28(16.9) | 1 |
| Health facility | 158(83.1) | 158(83.1) | 1 |
| **Wish to terminate the pregnancy earlier** |  |  |  |
| Yes | 142(85.5) | 108(65.1) | 0.0000001 |
| No | 24(14.5) | 58(34.9) | 0000001 |
| **Time taken to make decision after pregnancy confirmation** |  |  |  |
| Immediately | 115(69.3) | 162(97.6) | 0000001 |
| One month and above | 51(30.7) | 4(2.4) | 0000001 |
| **Reasons for delay in making the decision** | n=51 | n=4 |  |
| In a big dilemma whether to terminate or continue the pregnancy | 13(7.8) | 1(25) |  |
| Afraid to tell about my pregnancy | 6(3.6) | 3(74) |  |
| Thought of pregnancy termination is difficult | 29(17.5) |  |  |
| Assumed the pregnancy is too late | 3(1.8) |  |  |
| Religious reason | 4(2.4) |  |  |
| Moral issue | 3(1.8) |  |  |
| Thought my partner would help me to raise | 7(4.2) |  |  |

|  |  |  |
| --- | --- | --- |
| the baby |  |  |
| Concern on the condition and money required for pregnancy termination | 67(41.4) |  |
| Separated with my partner | 5(3) |  |
| Unprecedented event happens in my life | 25(15.1) |  |
| Other(I come immediately, I was in another area, I was too young) | 4(2.4) |  |
| **The period between your decision to terminate and the termination in weeks** |  |  |
| Mean(SD) in weeks. | 3.57(8.6) | 1.82(3.76) |
| **Reason for the delay to seek abortion care after a decision** |  |  |
| Lack of money to terminate the pregnancy | 41(24.7) | 109(65.7) |
| Don't have money for transportation | 23(13.9) | 1(0.6) |
| Being afraid to tell about my pregnancy | 46(27.7) | 8(4.8) |
| Did not know the place to get the service | 51(30.7) | 7(4.2) |
| Opposition from partner to terminate | 2(1.2) | 13(7.8) |
| Pressure from family/friends not terminate | 4(2.4) | 4(2.4) |
| Did not have time to terminate | 30(18.1) | 12(7.2) |
| Did not know earlier termination is better | 50(30.1) | 1(0.6) |
| Did not know about abortion service | 52(31.3) | 2(1.2) |
| Because of COVID 19 I did not visit HF | - | 5(3) |
| Other** | 5(3) | 6(5.6) |

\*Being in another area, I was too young;\*\*Health facility was closed,

### Respondents’ knowledge and belief on abortion service

A considerable number of respondents in the pre-intervention (62%) and post-intervention (43.4%) perceived that the communities understand/considered abortion as a sin. Seventy-seven percent of the respondents in the pre-intervention and 96.8% in the post-intervention believe that the community belief about abortion affect them to terminate their last pregnancy. The proportion of women who knew about the Ethiopian abortion law was increased from 26.5% in the pre-intervention to 81.9% in the post-intervention. The most common circumstance for legal abortion in the pre-intervention was rape (90.1%), incest (77.2%) and underage pregnancy (25%) while this number was 100% for rape,73.5% for incest and 54.2% for underage pregnancy in the post intervention period. The number of respondents who respond that safe abortion service is provided for free in a public health facilities was increased from 27.7% in the pre-intervention to 85.5% in the post-intervention period. Based on the post intervention data, 71.8% of the respondent’s report that they have heard about safe abortion service is provided from health facilities when they visited health facilities for a pregnancy test, they were informed by health professionals or provided them the paper while 24.6% of the respondents reported that they read from a poster when they visited health facilities for pregnancy test (Table 4).

**Table 4:** Knowledge of client about abortion service in health facilities of Mekelle City, Tigray region, Ethiopia, 2020.

| **Variable** | **Pre-intervention** | **Post intervention** | **P-value** |
| --- | --- | --- | --- |
|  | n(%) | n(%) |  |
| **How does the community understand abortion** |  |  |  |
| Considered as sin | 103(62) | 72(43.4) | 0.0006 |
| Promiscuousness | 46(27.7) | 23(13.9) | 0.0018 |
| Not sure | 17(10.2) | 71(42.8) | 0.00000001 |
| **Does the community beliefs affect you to terminate** | n=149 | n=95 |  |
| Yes | 128(77.1) | 92(96.8) | 0.0051 |
| No | 21(12.7) | 3(3.2) | 0.0051 |
| **Which of the things affect you** | n=128 | [n=95] |  |
| Considered as sin | 79(47.6) | [n=92] | 0.000000001 |
| Promiscuousness | 48(28.9) | 71(77.2) | 0.000000001 |
| **Know about Ethiopian abortion law?** |  |  |  |
| Yes | 44 (26.5) | 136(81.9) | 0.000000001 |
| No | 122 (73.5) | 30 (18.1) | 0.000000001 |
| **Circumstance to legal abortion service** | n=44 | n=136 |  |
| Rape | 40(90.1) | 136(100.0) | 0.00037 |
| Incest | 34 (77.2) | 100(73.5) | 0.620 |
| Lethal congenital anomalies of the fetus | 2(4.5) | 95 (69.8) | 0.000000001 |
| Mother has physical or mental disability | 2(4.5) | 89 (65.4) | 0.000000001 |
| Underage pregnancy | 11(25.0) | 90(54.2) | 0.000000001 |
| **Safe abortion provided in public health facilities for free** |  |  |  |
| Yes | 46 (27.7) | 142 (85.5) | 0.000000001 |
| No | 3 (1.8) | 3(1.8) | 1 |
| I don't know | 117 (70.5) | 21 (12.7) | 0.000000001 |
| **Source of information** | 46 | 142 |  |
| Informed by the health professionals/gave me a paper while visited health facilities for | - | 102 (71.8) | - |
| pregnancy test |  |  |  |
| Read a poster while visited health facilities for pregnancy test | - | 35 (24.6) | - |
| From friends, neighbors, and family | 46 (100) | 57(34.3) | 0.19 |

### Effect of the intervention in reducing the gestational age in weeks in termination of pregnancy

The mean of gestational age in weeks using ultrasound for the respondents in the pre-intervention group was 12.1(95% CI:11.18 to 13.05) as opposed to 7.5%(95% CI:7.14 to 7.85) in the post intervention group. Compared to the pre-intervention group, a much change in reducing the gestational age in weeks was observed in the post intervention period to terminate the pregnancy with 9.8 decrease per 100 respondents (95% CI 9.25 to 10.36) (Table 5). Along with a Cohen’s d with value of 5.23 was found (Table 5).

**Table 5:**
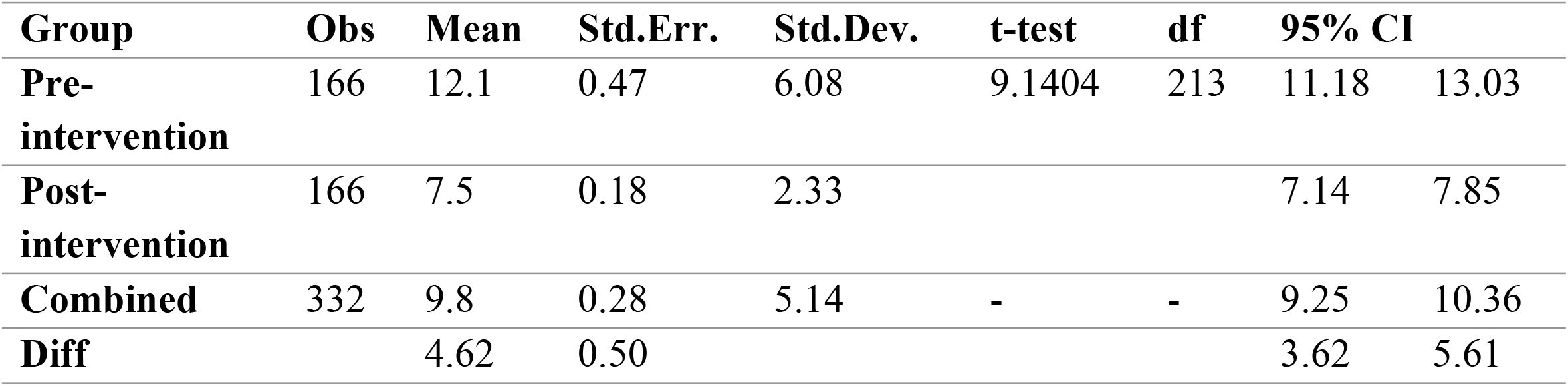
Estimated independent t-test coefficients to show the effect of the intervention in reducing the gestational age in weeks in termination of pregnancy in Mekelle City health facilities, Tigray region, Ethiopia, 2020

| Group | Obs | Mean | Std.Err. | Std.Dev. | t-test | df | 95% CI |  |
| --- | --- | --- | --- | --- | --- | --- | --- | --- |
| **Pre-intervention** | 166 | 12.1 | 0.47 | 6.08 | 9.1404 | 213 | 11.18 | 13.03 |
| **Post-intervention** | 166 | 7.5 | 0.18 | 2.33 |  |  | 7.14 | 7.85 |
| **Combined** | 332 | 9.8 | 0.28 | 5.14 | - | - | 9.25 | 10.36 |
| **Diff** |  | 4.62 | 0.50 |  |  |  | 3.62 | 5.61 |

### McNemar’s test for the knowledge of respondents on the possibility of termination pregnancy based on wish according to Ethiopia abortion law

Based on the table below, there was statistically significant difference between the pre and post-intervention on the respondent knowledge on the possibility of termination pregnancy based on wish according to Ethiopian abortion law at a p-value of <0.024(Table 6).

**Table 6:**
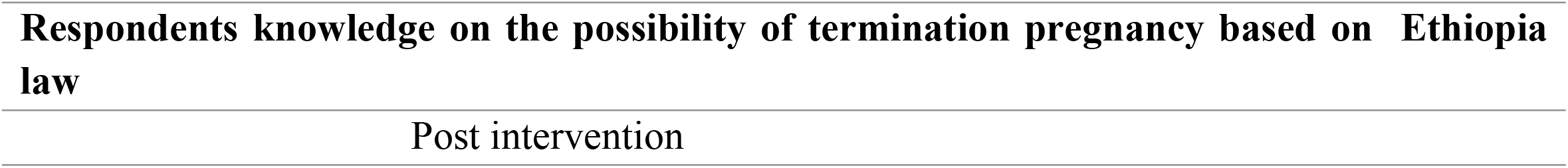
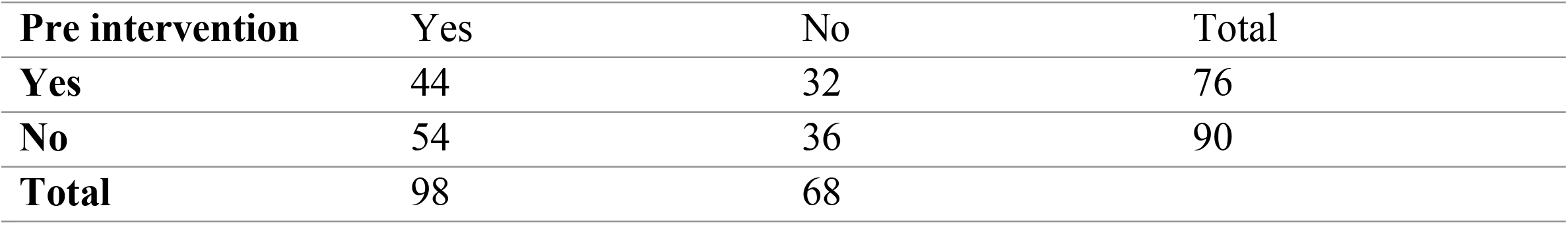
McNemar’s test for the knowledge of respondents on the possibility to terminate a pregnancy based on wish according to Ethiopia law in Mekelle City, Tigray region, Ethiopia, 2020

| **Pre intervention** | **Yes** | **No** | **Total** |
| --- | --- | --- | --- |
| **Yes** | 44 | 32 | 76 |
| **No** | 54 | 36 | 90 |
| **Total** | 98 | 68 |  |

## Discussion

The mean gestational age at termination of pregnancy was 12.12 (+6.23) in the pre-intervention while it was reduced to 7.5 (+2.33) weeks in the post-intervention. Along with there was a statistical significant difference in the women’s knowledge on the possibility of termination pregnancy according to Ethiopian abortion law at a p-value of <0.024

This study revealed that the intervention had a significant effect in the reduction of gestational age for termination pregnancy following the provision of street poster and a client information sheet. In this finding, the mean difference of the gestational age at termination of pregnancy showed a significant difference between pre and post intervention. Along with, Cohen’s d was calculated for the mean of gestational age to determine the effect size. Hence, it was found with value of d=5.26. This implies that strength of the effect size was large. The strength of the effect size can be determined as small for d=0.2, medium for d=0.5 and large. This implies that, the intervention brings a large effect size in reducing the gestational age for the termination of pregnancy.^14^

Consistent with our finding, previous study has shown that women’s delay in seeking an abortion service is apparent difficulty in accessing abortion services in early pregnancy.^15,16^ The factors associated with women’s decisions to seek timely care can be postulated at various levels; delay to suspect pregnancy, delay to decide to terminate the pregnancy, and delay to get abortion care.^14^ Proof of this, this study found that proportion of women with delay to make decisions after pregnancy confirmation was reduced in the post-intervention group (30.7% to 2.4 %). Furthermore, the mean stay in weeks was reduced from 3.57 weeks in the pre-intervention to 1.82 weeks in the post-intervention group. The commonest reasons for the delay to make a decision after pregnancy termination were thought of pregnancy termination difficulty (17.5%), and concern on the condition and money required for pregnancy termination (41%).^17^ This is consistent with a secondary analysis done by D.G. Foster et al, which states financial constraint and difficulty in making decision as culprits for delay in decision following pregnancy diagnosis.^18^Furthermore, a study done in Mexico corroborates that perceived difficulty of abortion care limits earlier decision for termination of pregnancy.^19^

Similarly, a prospective study done by Kebede et al indicated that lack of information were to get the service (AOR =3.907, CI 1.532–9.965) were variables associated with a presentation in second-trimester for termination of pregnancy.^20^ Multicenter qualitative study done in Ethiopia suggested that women were pushing pregnancies in to second trimester as they were not having information that abortion services are provided for free.^4^ Overall, this study confirms the multi-dimensional nature of factors that account for the delay in seeking an abortion, and factors relating to abortion service delivery. To this end, this study intervened at the second delay.

The study findings signal that most women in the post-intervention group (65.1%) would have preferred to have had their abortions earlier than they did. This is considerably higher in the pre-intervention group (85.5%). This is consistent with a study done in Mexico which reported a majority of the study participants wished if they have had their abortions earlier. Eighty-nine percent of participants who had pregnancies at 11–12 weeks of gestation when they contacted health indicated that they would have preferred to have obtained their abortion earlier, compared to 35% of those 7–10 weeks of gestation.^21^

We were not able to decrease this significantly because there were multiple players beyond the sphere of the intervention. Our study revealed that there was statistically significant difference between the pre- and post-intervention on respondent knowledge on the possibility of termination pregnancy. Similarly, other studies done in Amhara, Harar and Oromia Regions of Ethiopia reported that women who had a limited previous knowledge of the legal context of abortion and where to obtain an abortion service had led to delay in seeking abortion.^22-24^

To this end, the intervention geared at improving awareness of women that abortion services are provided at public health facilities for free and according to abortion law has decreased the contribution of these two in a delay to make a decision to terminate a pregnancy. This implies that creating awareness on the legalization of abortion based on the circumstance (rape, incest, lethal congenital anomalies of the fetus, mother has physical or mental disability and underage pregnancy) would have a contribution in reducing the consequence that results from delay in seeking abortion service.

The finding from the current study could be generalized to women who seek abortion service and it employed a pre-post study design with distributions of street poster and client information sheet to reduce the gestational age in seeking abortion service. The interviews were conducted by someone who is not involved in patient management. Thus, leading to decreased bias. However, the study has the following limitation. 1) It doesn’t employ an intended intervention before like advertisement of the health education message through radio or TV; 2) This doesn’t adjust the effect of the covariate or pre intervention (baseline) data in the final model as a result of using independent t-test and 3) It doesn’t use advanced study design like cluster randomized controlled trial (cRCT) to randomized group in to intervention and control group.

## Conclusion

The present study suggest that providing intervention on the availability of safe service and optimal gestational age for termination pregnancy has delay in seeking the abortion service with large effect size. In addition, a significant number of women increased their knowledge on the termination of pregnancy based on the national guide line. We recommend further rigorous study with strong design like the cluster randomized controlled trial by involving the three delay faces of safe abortion service would be vital.

## Data Availability

All data produced in the present work are contained in the manuscript

## Acknowledgments

We are grateful for the Family planning by choice project for generously funding the study. We would also like to thank all women who participated in this study for their cooperation in taking part in this study.

## Contributors

**AY** was involved in the conception, study design, execution, analysis, interpretation, and manuscript development. **MW** was involved in the conception, study design, data acquisition, and interpretation. **MA** was involved in study design, execution, analysis, interpretation, and manuscript write-up. **MH, AH, MT, BM, DG, HT** were involved in study design, data acquisition, and manuscript write-up. All authors read and approved the final manuscript.

## Data availability

The datasets generated and/or analyzed during the current study are not publicly available but are available from the corresponding author on reasonable request.

## Ethical statements

### Patient consent for publication

Consent obtained from patients, parent(s)/guardian(s).

### Ethics approval

This study involves human participants and was approved by the Institutional Review Board (IRB) of Mekelle University College of Health Sciences with a reference number of ERC-13159/2019

## References

1. Who.int. 2022. Abortion care guideline. [online] Available at: <https://www.who.int/publications/i/item/9789240039483> [Accessed 21 September 2022].

2. E-library.moh.gov.et. 2014. Technical and procedural guidelines for save abortion services in Ethiopia. [online] Available at: <https://e-library.moh.gov.et/library/wp-content/uploads/2021/07/SAC-guidline-English-June-2014-1.pdf> [Accessed 21 September 2022].

3. Lalitkumar S, Bygdeman M, Gemzell-Danielsson K. Mid-trimester induced abortion: a review. Human Reproduction Update. 2007;13(1):37–52.

4. Ahmed Abdela, Tibebu Alemayehu, Takele Geressu, Yirgu Gebrehiwot, Alison Edelman, Second trimester abortion: current practices and barriers to services in Ethiopia, 2010, Ipas Ethiopia

5. Edelman, A., Alemayehu, T., Gebrehiwot, Y., Kidenemariam, S. and Getachew, Y., 2014. Addressing unmet need by expanding access to safe second trimester medical abortion services in Ethiopia, 2010 – 2014. International Journal of Gynecology &amp; Obstetrics, 128(2), pp.177–178.

6. Ahman SE. Unsafe abortion: global and regional incidence, trends, consequences, and challenges. Journal ofObstetrics and Gynaecology Canada. 2009;31(12):1149–58.

7. Sedgh G, Singh S, Shah IH, Åhman E, Henshaw SK, Bankole A. Induced abortion: incidence and trends worldwide from 1995 to 2008. The Lancet. 2012;379(9816):625–32.

8. Facts on Unintended Pregnancy and Abortion in Ethiopia, 2010 [Internet]. Guttmacher Institute; 2010 [cited February 17. 2019]. Available from: https://info@guttmacher.org.

9. Unsafe Abortion. Global and Regional Estimates of the Incidence of Unsafe Abortion and Associated Mortality in 2003. Geneva: WHO. 2007.

10. ICF. CSACEa. Ethiopia Demographic and Health Survey. Addis Ababa, Ethiopia, and Rockville, Maryland, USA: CSA and ICF. 2016.

11. Legesse TW, Solomon TM, Teresa KB. Unsafe abortion and associated factors among women in reproductive age group in Arsi Zone, Central Ethiopia. International Journal of Nursing and Midwifery. 2017;9(10):121–8.

12. Siraneh YW A. Determinants and Outcome of Safe Second Trimester Medical Abortion at Jimma University Medical Center, Southwest Ethiopia. Journal of pregnancy. 2019;2019:4513827.

13. Mulat A, Bayu H, Mellie H, Alemu A. Induced second trimester abortion and associated factors in Amhara region referral hospitals. BioMed research international. 2015;2015:256534.

14. McLeod S. McLeod S. What does effect size tell you? Simply psychology: 2019; 2–5.Available from: https://www.simplypsychology.org/effect-size.html.

15. Barnes-Josiah D, Myntti C, Augustin A. The “three delays” as a framework for examining maternal mortality in Haiti. Social Science & Medicine. 1998;46(8):981–93

16. Drey EA, Foster DG, Jackson RA,et al. Risk factors associated with presenting for abortion in the second trimester. Obstetrics and Gynecolgy 2006; 107(1):128–35

17. Finner L, Frohwirth LF, Dauphinee LA, et al. Timming of steps and the reasons for delays in obtaining abortions in the United States. Contraception 2006;74:334–44

18. D.G. Foster et al. / Contraception 77 (2008) 289–293. Foster D, Jackson R, Cosby K, Weitz T, Darney P, Drey E. Predictors of delay in each step leading to an abortion. Contraception. 2008;77(4):289–293.

19. Camille Garnsey, Alexandra Wollum, SofíaGarduño Huerta, OrianaLópez Uribe, Brianna Keefe-Oates & Sarah E. Baum (2022) Factors influencing abortion decisions, delays, and experiences with abortion accompaniment in Mexico among women living outside Mexico City: results from a cross-sectional study, Sexual and Reproductive Health Matters, 29:3, 2038359, DOI: 10.1080/26410397.2022.2038359

20. Kebede K, Gashawbeza B, Gebremedhin S, Tolu L. Magnitude and Determinants of the Late Request for Safe Abortion Care Among Women Seeking Abortion Care at a Tertiary Referral Hospital in Ethiopia: A Cross-Sectional Study. International Journal of Women’s Health. 2021; Volume 12:1223–1231.

21. Garnsey C, Wollum A, Garduño Huerta S, Uribe O, Keefe-Oates B, Baum S. Factors influencing abortion decisions, delays, and experiences with abortion accompaniment in Mexico among women living outside Mexico City: results from a cross-sectional study. Sexual and Reproductive Health Matters. 2022;29(3).

22. Geleto A, Markos J. Awareness of female students attending higher educational institutions toward legalization of safe abortion and associated factors, Harari Region, Eastern Ethiopia: a cross sectional study. Reproductive Health. 2015;12(1).

23. Seid, H. Yeneneh, B. Sende, and S. Belete, “Barriers to accessing safe abortion services in East Shoa and Arsi zones of Oromia regional state, Ethiopia,” The Ethiopian Journal of Health Development (EJHD), vol. 29, no. 1, 2016.

24. Wasihun Y, Mekonnen T, Asrat A, Dagne S, Menber Y, Fentahun N. Determinants of Second-Trimester Safe Termination of Pregnancy in Public Health Facilities of Amhara Region, Northwest Ethiopia: An Unmatched Case-Control Study. Advances in Public Health. 2021;2021:1–7.

